# Inflammasome-related markers at the ICU admission are not associated with outcome in the critically ill COVID-19 patients

**DOI:** 10.1101/2021.10.30.21265662

**Authors:** Barbara Adamik, Magdalena Ambrożek-Latecka, Barbara Dragan, Aldona Jeznach, Jakub Śmiechowicz, Waldemar Goździk, Tomasz Skirecki

**Affiliations:** Department of the Anaesthesiology and Intensive Therapy, Wroclaw Medical University, Wroclaw, Poland; Department of Clinical Cytology, Centre of Postgraduate Medical Education, Warsaw, Poland; Laboratory of Flow Cytometry, Centre of Postgraduate Medical Education, Warsaw, Poland

**Keywords:** SARS-CoV2, biomarker, inflammation, caspase-1, critical care, interleukin-1

## Abstract

**Purpose:** Development of targeted biological therapies for COVID-19 requires reliable biomarkers that could help indicate the responding patients. Hyperactivation of the inflammasome by SARS-CoV2 virus is hypothesized to contribute to severe course of the COVID-19 disease. Therefore, we aimed to evaluate the prognostic value of several inflammasome-related cytokines and proteins at the admission to the intensive care unit (ICU).

**Patients and methods:** Plasma samples were obtained from 45 critically ill COVID-19 patients and from 10 patients any without any signs of infection (TBI, traumatic brain injury) on admission to the ICU. The concentration of IL-1*α*, IL-1*β*, IL-18, IL-1RA, galectin-1, ASC, LDH, ferritin, and gasdermin D were analyzed. A novel cell-free caspase-1 plasma assay was developed by inhibitor-based immunoprecipitation followed by Western Blot. Demographic and clinical characteristics were recorded.

**Results:** In-hospital mortality in COVID-19 patients reached 62%. Galectin-1 was 1.8-fold lower in COVID-19 than in TBI patients (17101.84 vs. 30764.20 pg/ml, p=0.007), but other inflammasome-related biomarkers were at similar concentrations. Patients with SOFA score of >9 on admission who were at high risk of death had significantly higher galectin-1 but lower IL-1RA in comparison to low-risk patients (25551.3 pg/ml vs 16302.7 pg/ml, p=0.014; 14.5 pg/ml vs 39.4 pg/ml, p=0.04, respectively). Statistically significant correlations were observed between: IL-1*α* and platelets (r=-0.37), IL-1*β* and platelets (r=-0.36), ferritin and INR (r=0.39). Activated caspase-1 p35 was detectable in 12/22 COVID-19 patients but in none of the TBI patients. Its presence was related with higher fibrinogen and lower D-dimers. Moreover, the densitometric analysis showed a significantly higher amount of p35 in patients with SOFA> 9.

**Conclusion:** Our results indicate that the systemic markers of activation of the inflammasome in critically ill COVID-19 patients is not directly related with outcome. Therefore, potential interventions aimed at the inflammasome pathway in this group of patients may be of limited effectiveness and should be biomarker-guided.

## Introduction

Infection with the respiratory SARS-CoV2 virus causes the coronavirus disease 2019 (COVID-19) which ranges from asymptomatic infection through mild and severe disease (pneumonia) to critical disease which has a form of the acute respiratory distress syndrome (ARDS) or sepsis and septic shock^1^. This range of severity is reflected by the mortality ranges which reaches approx. 40% in critically ill patients^2^. Although other coronavirues can infect humans, the COVID-19 is characterized by unique pathogenesis making it a distinct disease^3^. Aside from respiratory support, the only widely accepted treatment for COVID-19 is dexamethasone and recently tocilizumab^1^.

There is an urgent need to develop efficient therapies for COVID-19 and it becomes apparent that as in the case of bacterial sepsis, a biomarker-guided personalized approach is a prerequisite for successful biological therapies^4^. Since early pandemics we among others have proposed that the unbalanced inflammasome signaling may be a key process in the pathogenesis of COVID-19^5,6^. Briefly, inflammasomes are multiprotein complexes activated by various stimuli such as dsDNA (Absent in Melanoma, AIM2), potassium efflux and reactive oxygen species (NLR Family Pyrin Domain Containing 3, NLRP3)^7^. Upon activation, the receptor proteins oligomerize and recruit adaptor proteins (apoptosis-associated speck-like protein, ASC) and pro-caspase-1. Then pro-caspase-1 undergoes autoproteolysis to its active form which then can cleave pro-interleukin-1*α, β*, -18 and gasdermin D (GSDMD). N-terminal GSDMD can form pores in the cell membrane which leads to the release of the cleaved cytokines as well as other molecules (such as lactate dehydrogenase; alarmins) and lytic cell death called pyroptosis^7^. Indeed, it has been demonstrated that the inflammasome is activated in the lungs of COVID-19 patients^8,9^ and can be activated in the circulating monocytes^10^. However, the status of inflammasome reactivity in the blood myeloid cells appears complexed^11^. Although some studies showed an increase in concentrations of pro-inflammatory cytokines with increasing severity of COVID-19^12^, there is a limited number of studies focused on critically ill COVID-19 patients who are at the highest risk of death. Nevertheless, there are ongoing clinical trials with direct (Dapansutrile, Colchicine) inflammasome inhibitors and indirect (Disulfiram, a GSDMD inhibitor). Also, recently two studies on the IL-1R antagonist in COVID-19 were published with opposite results^13,14^.

In this study, we aimed to evaluate the prognostic utility of selected plasma proteins related to inflammasome activation in critically ill COVID-19 patients. Considering the high mortality and lack of effective therapies we focused on the critically ill COVID-19 patients at the admission to intensive care unit (ICU). We hypothesized that biomarkers of good accuracy could serve to guide immunomodulatory therapies targeting the inflammasomes.

## Material and methods

### Patients

We have prospectively collected samples from 45 patients with critical COVID-19 at the admission to ICU of the Wroclaw Medical University. Critical COVID-19 was defined according to COVID-19 treatment guidelines^1^. Briefly, the patients filled criteria for acute respiratory distress syndrome (ARDS), sepsis, septic shock, and required introduction of life-sustaining therapies^1^. SARS-CoV2 infection was confirmed by real-time reverse-transcriptase polymerase chain reaction (RT-PCR) assay of nasal or pharyngeal swab probes. All patients were treated according to the WHO guidelines^1^. All patients were administered intravenous dexamethasone at a dose of 6 mg per day. The study protocol complies with the 1975 Declaration of Helsinki as revised in 1983. The Wroclaw Medical University Bioethics Committee approved this study (consent No. 394/2021). The Committee also approved the collection of plasma from the traumatic brain injury (TBI) (consent No.KB-391/2015). Written informed consent was obtained from the patient or a legally authorized representative. Blood samples collected from patients with TBI were used to compare markers of inflammasome activation between cases with SARS-CoV2 infection (COVID-19 group) and cases without any signs of infection (TBI group). The clinical status of the patients was determined with the Acute Physiology and Chronic Health Evaluation (APACHE) II score and Sequential Organ Failure Assessment (SOFA) score on admission to the ICU. The APACHE II score it is routinely used as a prediction tool for ICU patients and includes 12 physiological variables (the fraction of inspired oxygen, partial pressure of oxygen, body temperature, mean arterial pressure, blood pH, heart rate, respiratory rate, serum sodium, serum potassium, serum creatinine, hematocrit, white blood cell count, and the Glasgow Coma Scale) and 2 disease-related variables (the history of severe organ failure or immunocompromised and the type of ICU admission). The SOFA score is routinely used in the ICU for monitoring the severity of patient’s clinical condition based on the status of the following systems: respiratory (PaO2/FiO2 index), cardiovascular (mean arterial pressure and the dose of vasopressors), hepatic (bilirubin level), coagulation (platelets level), renal (creatinine level/urine output), and neurological (Glasgow coma scale).

### Cytokine analysis

The blood samples were collected via routinely inserted arterial cannula. 2.7 ml of peripheral blood was collected into 0,109 M sodium citrate containing tubes (BD Vacutainer, BD, NJ, USA). Plasma was collected after centrifugation at 2000 x g for 10 minutes and stored at -70°C. IL-1*α*, IL-1*β*, IL-18, IL-1RA and galectin-1 concentrations were analyzed by Milliplex Human Multiplex Assay (Merck, Germany) using the Magpix System. ACS protein was analyzed using the commercially available ELISA kit (Cusabio Technology, TX, USA). Human GSDMD was analyzed using commercial ELISA kit (Abclonal, MA, USA). Other analytes were measured at the certified hospital clinical laboratory.

### Caspase-1 immunoprecipitation

Equal volumes of plasma (200 µl) from Controls (TBI) and COVID-19 patients were treated with 50 µM Biotin-FAD-FMK (SantaCruz Biotechnology, CA, USA) with gentle rotation at 4°C overnight. Then, 10 µl of Streptavidin Sepharose Bead Conjugate (Cell Signalling Technology, MA, USA) was added to plasma samples and incubated for 5h with gentle rotation at 4°C. The biotin-streptavidin bead complexes were centrifuged for 5 min at 2300 x g and washed 5 times using 1 x Ripa Lysis buffer (Millipore) supplemented with Protease Inhibitor Cocktail (Millipore) and 1 mM PMSF. Pellet samples were suspended in 4 x Laemmli buffer and boiled for 3 min in 94°C. For detection of the active forms’ caspase-1 the pulled down immunocomplexes were separated in 15 % SDS - PAGE gels following by wet electrotransfer onto PVDF membranes. Non-specific bindings were blocked in TBS-T with 5 % non – fat milk for 1 h in room temperature. Membranes were incubated with anti – Caspase-1 (cleaved 297; 1:1000 dilution; GenTex) overnight at 4°C. Rabbit polyclonal antibody (1: 10000 dilution; Vector Laboratories, Inc.) was used to detect primary antibody. Signals were visualized by WesternBright^*®*^ ECL HRP substrate (Advansta) using UVITEC gel documentation system. Densitometric analysis was done by GelAnalyzer 19.1.

### Statistical analysis

Normality of data was tested using the Shapiro-Wilk test. Continuous variables are expressed as median values and interquartile ranges whereas categorical variables are presented as frequencies with percentages. Categorical variables were compared using the Fisher’s exact test. Continuous variables were compared using the Mann-Whitney test. Receiver operating characteristic (ROC) curves were constructed to calculate area under the curve (AUC) of the investigated parameters. The cut-off values were indicated using the Youden’s index method. Correlations between variables were tested using the Spearman’s test. Logistic regression analysis was performed to test the relationship between biomarkers and the risk of death or development of secondary infections. Statistica 13.1 software was used for calculations and GraphpadPrism 9.0 was used to create graphs. Statistical significance was determined as p < 0.05.

## Results

### Clinical characteristics of the patient’s population

We have enrolled forty-five critically ill COVID-19 patients with RT-PCR confirmed SARS-CoV2 infection. The clinical and demographic characteristic of the group is presented in **Tab. 1**. This group included 24 males (53%) and 21 females and the median age was 58 years (IQ: 43-68). At admission, the patients’ median APACHE II score was 14 (IQ: 11-22) and SOFA score was 9 (IQ:7-10). The hospital mortality reached 62% and was higher than 28-day mortality which was 53%. The patients who did not survive were significantly older (median 64 (IQ:52-70) vs. 50 (IQ: 33-60) yrs.) and had higher APACHE II score in comparison to the survivors (20 (IQ:14-25) vs. 12 (IQ:8-13)). Most common comorbidities were hypertension and cancer; however, their occurrence was not related with survival. The extra corporeal membrane oxygenation (ECMO) therapy was used in 29% of non-survivors and 18% of survivors. Twelve patients were admitted with co-infections with *Acinetobacter baumannii* and *Candida albicans* being the most common pathogens (3/12 each) followed by the methicillin-resistant *Staphylococcus aureus* (MRSA, 2/12). Twenty-one patients (47%) developed secondary infections with *A. baumannii* and *C. albicans* as the most frequent pathogens (6/21 each) followed by MRSA and *Stenotrophomonas maltophilia* (4/21 each).

### Markers of inflammasome activation are not outcome-related in critically ill COVID-19 patients

Firstly, we have compared the levels of IL-1*α*, IL-1*β*, IL-18, IL-1RA, galectin-1, ASC and gasdermin D in the study group with the age and sex matched group of traumatic brain injury (TBI) patients. Interestingly, only galectin-1 was significantly different in the critically ill COVID-19 patients in comparison to TBI patients (17101.84 (IQ: 10836.62 - 25551.30) pg/ml vs. 30764.20 (IQ: 23150.67 - 35759.48) pg/ml; p=0.007; **Fig. 1**). Then, we compared the concentration of the inflammasome related markers between COVID-19 survivors and non-survivors. Neither of the proteins could differentiate between these groups (**Fig. 2)**. Moreover, LDH and ferritin which can serve as surrogate markers of cell death and IL-1 signaling, respectively, were not increased in patients with unfavorable outcome (**Tab. 2**). Other routinely monitored markers of inflammation as white blood count, IL-6 or CRP were also similar in these two groups. Of the analyzed parameters, only the international normalized ratio (INR) was significantly higher in the non-surviving COVID-19 patients. However, logistic regression analysis did not reveal the relationship between INR either other analyzed parameters and mortality (p>0.05). Also, there were no difference in the concentrations of the analyzed biomarkers of the inflammasome response between patients who were admitted with co-infections and those who were not. These results indicate that inflammasome-related proteins are less elevated in critically ill COVID-19 patients at ICU admission than in TBI patients and are not different between survivors and non-survivors.

**Figure 1.**
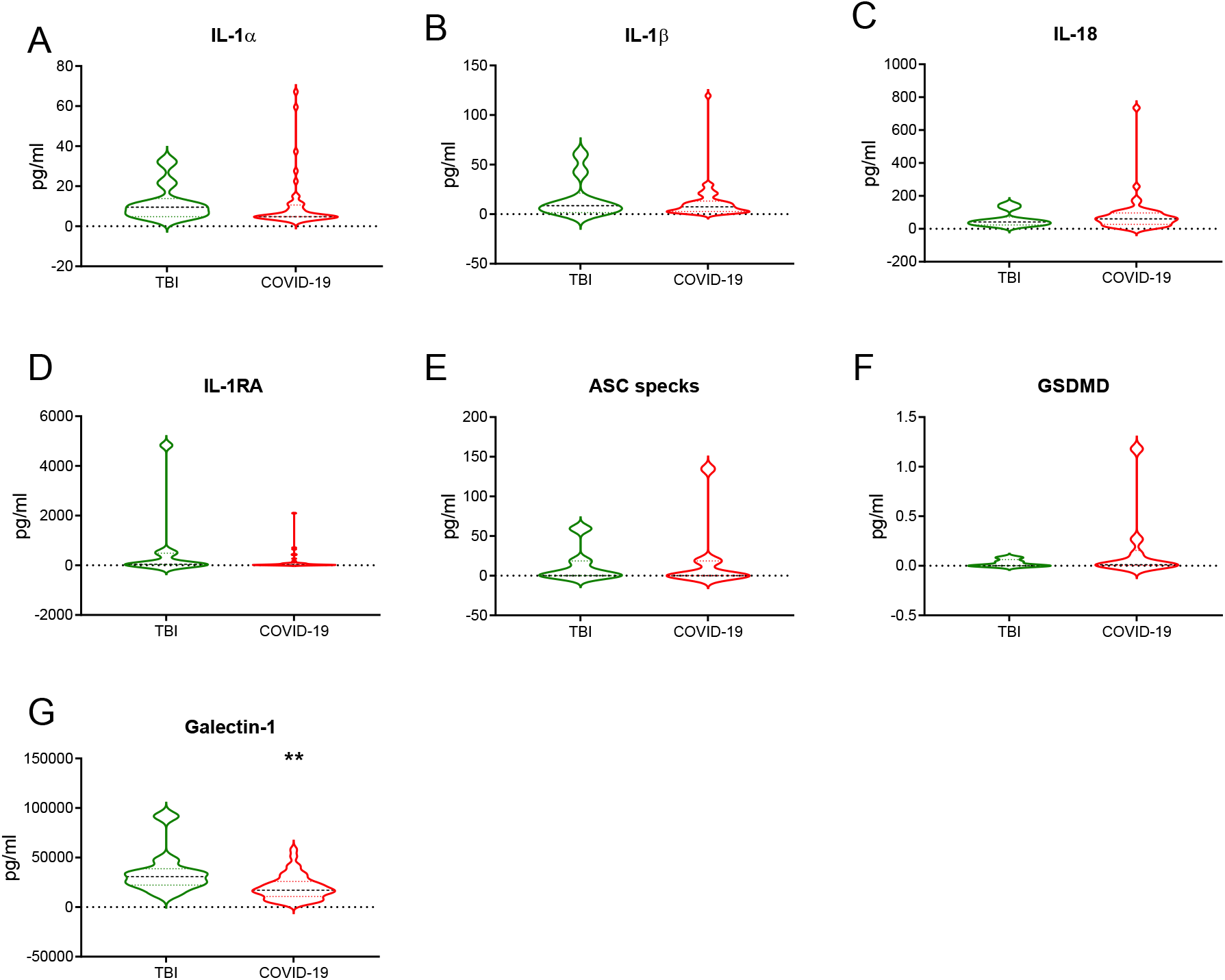
Comparison of the inflammasome-related markers in the traumatic brain injury (TBI) and critical COVID-19 patients. A. Interleukin-1*α*. B. Interleukin-1*β*. C. Interleukin-18. D. Interleukin-1 Receptor Antagonist. E. Apoptosis-associated speck-like protein containing a CARD. F. Gasdermin D. G. Galectin-1. Concentrations of plasma proteins were compared between TBI patients (n=10) with critically ill COVID-19 (n=45) with the Mann-Whitney test. **p<0.05.

**Figure 2.**
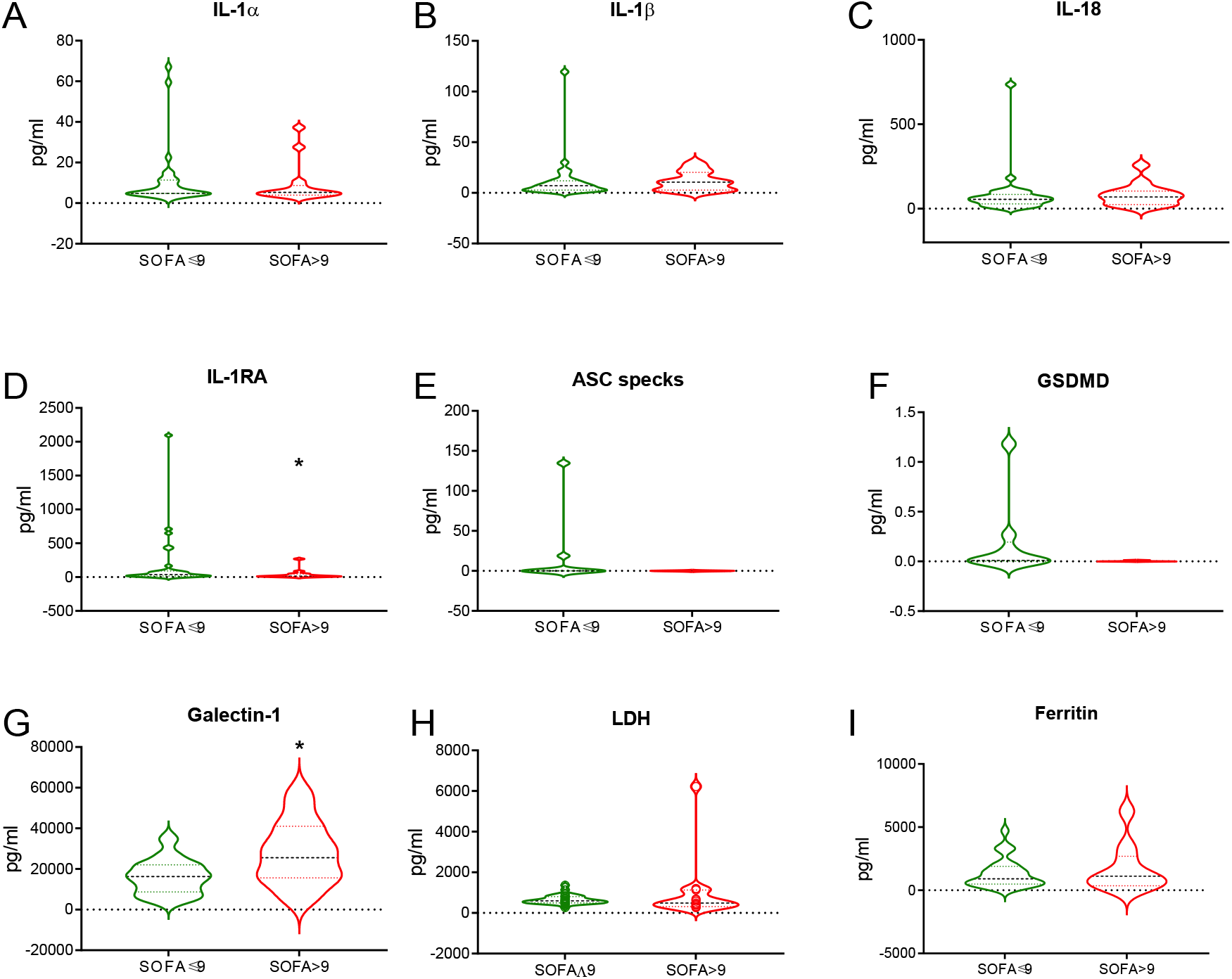
Inflammasome-related markers in the plasma of critically ill COVID-19 patients at low- and high risk of death. The high risk of death was stratified by the threshold of more than 9 points in SOFA score. A. Interleukin-1*α*. B. Interleukin-1*β*. C. Interleukin-18. D. Interleukin-1 Receptor Antagonist. E. Apoptosis-associated speck-like protein containing a CARD. F. Gasdermin D. G. Galectin-1. H. Lactate Dehydrogenase. I. Ferritin. Concentrations of plasma proteins were compared between patients at high risk (n=15) with patients at low risk of death (n=30) with the Mann-Whitney test. *p<0.05.

### Galectin-1 and IL-1RA can stratify the severity of critically ill COVID-19 at the admission

As the SOFA score is one of the most widely used tools to stratify the critically ill patients including COVID-19, we analyzed its predictive capacity in our group of patients. Indeed, the ROC analysis for in-hospital mortality was characterized by the area under the curve (AUC) of 0.824 (95%CI: 0.703 - 0.944) which is of moderate accuracy ^15^. As a predictor of death, the SOFA score of 9 points was characterized by the sensitivity of 75.0% and specificity of 64.7%. Then, we applied this cut-off point to divide the patients into two groups of high- and low-risk of death. The high-risk patients had significantly higher level of galectin-1 in comparison to individuals at low-risk (25551.30 (IQ: 15628.85 - 41048.02) pg/ml vs. 16302.72 (IQ: 8785.86 - 22040.25) pg/ml; p=0.014, respectively). Interestingly, the high-risk group had significantly lower level of IL-1RA in comparison to the low-risk patients (14.50 (IQ: 9.28 - 47.01) vs. 39.35 (IQ: 17.87 - 88.01) pg/ml; p=0.047). The high-risk group patients were also characterized by higher white blood count (17.00 (IQ: 13.40 - 20.30) ×10^6^/ml vs. 12.75 (IQ: 10.20 - 16.80) ×10^6^/ml; p=0.034), procalcitonin (2.10 (IQ: 0.40 - 4.00) ng/mL vs 0.25 (IQ: 0.11 - 0.60) ng/L; p=0.001 and IL-6 (700.00 (IQ: 680.70 - 1890.00) pg/ml vs. 66.60 (IQ: 16.00 - 107.00) pg/ml; p=0.001). The frequency of co-infections was not significantly different between these groups, however chronic comorbidities such as coronary artery disease, heart failure and renal disease were significantly more common in the high-risk patients. Of interest, patients who developed secondary infections had lower level of galectin-1 than those who did not (14908.51 (IQ: 8394.48 - 21,911.48) pg/ml vs. 23,839.62 (IQ: 15,628.85 - 29,846.66) pg/ml; p=0.039, however logistic regression analysis did not confirm this relationship (OR=1.00, p=0.042). There were no differences among analyzed inflammasome-related biomarkers either standard inflammatory parameters (IL-6, PCT, WBC) between patients who developed secondary infections during ICU stay and those who did not. Altogether, these results suggest that the threshold of 9 SOFA score points indicated low- and high-risk critically ill COVID-19 patients who differed in the inflammasome and inflammatory profile.

### Correlations of the inflammasome-related markers with other inflammatory markers in the critically ill COVID-19

Additionally, we tested whether the concentrations of the inflammasome-related proteins are related with other measures. The Spearman’s correlation test revealed a weak but significant correlation between IL-1*α* and IL-1*β* (r=0.40) and between IL-18 and IL-1RA (r=0.60) and galectin-1 (r=0.27). Gasdermin D correlated with the ASC protein (r=0.44) and ferritin correlated with INR (r=0.39) and LDH (r=0.45). There also inverse correlation between IL-1*α* and platelets and IL-1*β* and platelets count (r=-0.37 and r=-0.36, respectively). No significant correlations were found between the inflammasome-related markers and CRP, PCT, nor IL-6. These observations support the hypothesis that the plasma concentration of analyzed inflammasome-related proteins are concomitantly regulated and reflect the same cellular processes.

### Cell free caspase-1 is not ubiquitously present in the circulation of critical COVID-19 patients

As secretion of IL-1 and pyroptosis can result from the activation of other than inflammasome cellular pathways^16^ we decided to confirm the activation of inflammasome in the COVID-19 patients by development of inhibitor-based immunoprecipitation of active caspase-1 followed by Western Blot detection of the specific fragments. Interestingly, the dominant form of caspase-1 present in the plasma was the intermediate form p35 found in 12 of 22 COVID -19 patients (**Fig. 3A**) while it was undetectable in TBI patients. In six of 22 COVID-19 patients also mature p20 form could be detected (**Fig. 3A**). Presence of active caspase-1 was not related with outcome neither severity of the disease. There was also no difference in caspase-1 between patients with coinfections. However, patients at high-risk of death defined by SOFA score >9 had significantly higher amount of p35 in densitometric analysis (**Fig. 3B**) Of interest, patients with detectable p35 had higher fibrinogen level (7.30 g/L (IQ: 5.50 - 8.70) vs. 5.45 g/L (IQ: 4.10 - 6.30); p=0.020) and lower d-dimers (1.35 μg/mL (IQ: 1.20 - 4.35) vs. 10.25 μg/mL (IQ: 6.30 - 20.90); p=0.019).

**Figure 3.**
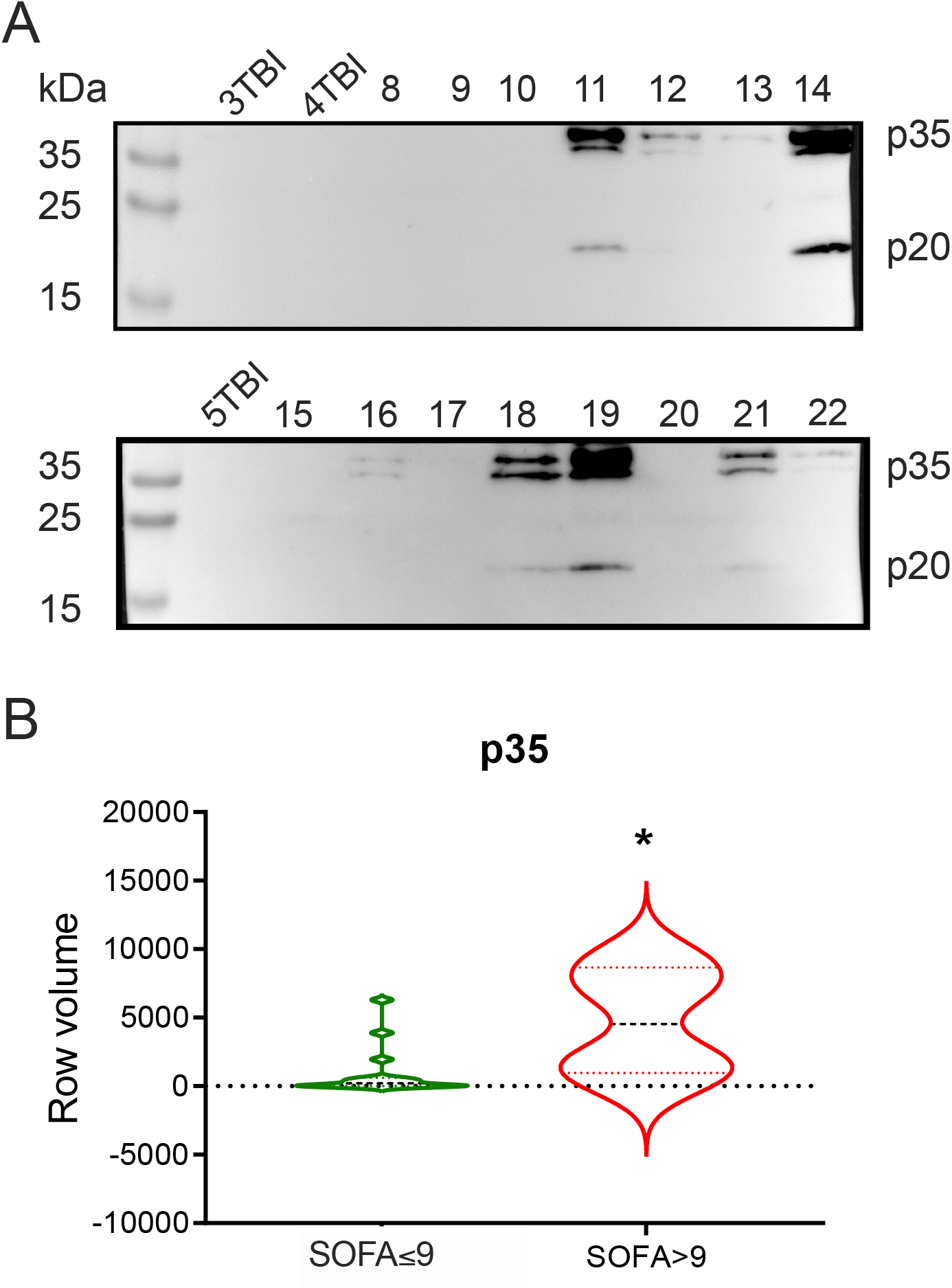
Active caspase-1 is unambiguously present in critical COVID-19 patient’s plasma. A. Representative immunoblots with labeled with anti-caspase-1 of the pull-downed FAD-FMK bonded caspases from the plasma of TBI patients (3TBI, 5 TBI) and COVID-19 patients (numbers 8-22). The mature p20 band is present in approx. half of the patients with intermediate p35 form.B. Patients at high risk of death (SOFA>9) had more circulating p35 as revealed by densitometric analysis. Data were compared with the Mann-Whitney test. *p<0.05.

## Discussion

Our study is the first to our best knowledge that have investigated comprehensively the plasma biomarkers related with inflammasome regulation in a well-defined group of critically ill COVID-19 patients at the ICU admission. We found that the marked activation of inflammasome defined as presence of active capsase-1 in the patient’s plasma is not ubiquitous and not related with outcome. Also, other quantitative inflammasome-related proteins and cytokines are not related with outcome and only galectin-1 and IL-1RA differed in patients with low- and high-risk of death.

In this study we have focused on the critically ill COVID-19 patients at admission to ICU as this group is at high risk of death (reflected by 28-day mortality of 53% outreached by in-hospital mortality of 62% in our group). Although many groups have tried to identify new potential biomarkers, we have selected a panel of proteins related with different stages of the inflammasome activity. It included the structural component of inflammasome, the ASC protein; effector proteins cleaved by caspase-1 (IL-1*α*, IL-1*β*, IL-18), their antagonist IL-1RA and gasdermin D which forms pyroptotic pores in cell membrane. Also, we measured galectin-1 and LDH which are markers of inflammatory cell death^17^ and ferritin which is induced by the inflammasome activity^18^. Moreover, we have established a new method to analyze active caspase-1 from human plasma which is based on the immunoprecipitation with biotinylated FAD-FMK, a pan-caspase inhibitor followed by Western Blot to specifically detected caspase-1 forms.

The concentration of the measures cytokines was not different between critical COVID-19 and TBI patients. We choose the TBI patients for comparison as it relatively homogenous group with sterile injury, age- and sex-matched. ASC, IL-1 and capase-1 were already shown to be upregulated in plasma of TBI patients ^19^. Although increased galectin-1 was already described in COVID-19 patients ^20^ it was compared to healthy controls only. Here, we found higher galectin-1 in TBI patients than COVID-19 which suggests that the latter is not characterized by extreme over-inflammatory response as suggested by some authors^3,21^. Our results expand the findings of others showing similar levels of inflammatory cytokines in COVID-19 and sepsis or other critical conditions ^22-24^.

Neither of the analyzed protein was related with mortality in critically ill COVID-19 patients. Although some of these cytokines like IL-18 or IL-1RA were already linked with the severity of COVID-19^9,25,26^ it should be highlighted that our study is focused on a defined population of critical COVID-19 in contrast to most studies which compare cytokine levels in early COVID-19 of various severity. As the analyzed cytokines can be produced in inflammasome-independent manner, we assessed the presence of active caspase-1. We have detected the intermediate p35 form of caspase-1 in 54% cases and mature p20 in 27% patients. These finding clearly indicate that level of inflammasome activation is heterogenous in critical COVID-19 and complete maturation of caspase-1 is not common in these patients. The status of circulating caspase-1 was not related with outcome similarly to the findings by the group of D. Zamboni who utilized capsase-1 ELISA^9^. Even less frequently we were able to detect the ASC and GSDMD proteins which also indicates infrequent activation of inflammasome in critical COVID-19. Our results stay in accordance with recent findings that showed only small percentage of NLRP3 and AIM2 inflammasome activation in the circulating monocytes from COVID-19 patients^10,27^. Moreover, current data point at monocytes and macrophages as the major lung cell types showing activation of the inflammasome in COVID-19^9,10^. Additionally, we compared inflammasome-related proteins in the patients divided by their SOFA score at admission with the most accurate threshold to predict mortality (low- and high-risk groups). Notably, galectin-1 and caspae-1 p35 was increased in the high-risk group, while IL-1RA was decreased. Such pattern may suggest indeed loss of control of this inflammatory pathway in the high-risk group of patients. The high-risk patients had also higher leukocyte count, IL-6, procalcitonin and INR however, neither of these parameters was significantly related with outcome in logistic regression model.

As inflammasomes are reported to play role in the susceptibility to secondary infections^28,29^ we tested for the relation between inflammasome activation markers and co-infections or development of secondary infections however, there were no significant correlations between these variables. We observed some weak but significant correlations between IL-1*α* and IL-1*β*, ASC and GSDMD, ferritin and LDH which suggests coordinated regulation of these mediators. Interestingly, we observed links between the inflammasome activity and dysregulated coagulation system. Patients with unfavorable prognosis had marks of coagulopathy indicated by higher INR values. In addition, there was a correlation between ferritin and INR and inversed correlation between IL-1*α* and IL-1*β* and platelets. Altogether, these observations suggest mechanistic link between inflammasome activation and coagulopathy and possibly features of secondary hemophagocytic lymphohistiocytosis^3^.

This study has several limitations related with small patients’ group and lack of mechanistic investigations. Noteworthy, our study group is well-defined and limited to the ARDS and sepsis forms of COVID-19. Although we analyzed several proteins related with activation of the inflammasome we did not measure other relevant mediators that can interact with this pathway (e.g. interferons). The present study would also benefit from the flow cytometry data of the inflammasome activation in circulating blood cells. Moreover, we analyzed only plasma proteins and it should be noted that they may not fully reflect the tissue response. Finally, we were not able to explain the heterogenic pattern of the inflammasome activity in these patients.

## Conclusion

Our findings are of important clinical relevance as inflammasome-targeted therapies are widely testes in COVID-19, mostly without the use of biomarker guidance. Recently, the negative results of the collaborative trial on the colchicine (which inhibits inflammasome formation^30^) were published^31^. Also, trials targeting IL-1 in severe COVID-19 with anakinra was reported to be negative^14^. At the same time, another study that used su-PAR levels to guide the anakinra treatment in early COVID-19 showed significant benefit^13^. The results of these studies are not surprising in the light of our findings showing heterogeneity in inflammasome activation in critically ill COVID-19 patients and lack of its direct relation with mortality. Yet, it could be speculated that in a selected subpopulation of patients with activated caspase-1 or high galectin-1 level these therapies could turn beneficial. Nevertheless, further mechanistic studies on the role of inflammasome in the pathogenesis of COVID-19 are needed.

## Data Availability

All data produced in the present study are available upon reasonable request to the author

## Authors contribution

All authors made substantial contributions to conception and design, acquisition of data, or analysis and interpretation of data; took part in drafting the article or revising it critically for important intellectual content; agreed to submit to the current journal; gave final approval of the version to be published; and agree to be accountable for all aspects of the work.

---

**Table 1.** Clinical and demographical characteristics of the critically ill COVID-19 patients

| Characteristic | survivors (n=17) | non-survivors<br>(n=28) | p |
| --- | --- | --- | --- |
| Age | 50 (33-60) | 64 (52-70) | **0.0030** |
| Sex (male), n (%) | 8 (47) | 16 (57) | 0.1248 |
| Hospitalization days | 28 (13-45) | 17 (7-28) | **0.0349** |
| ICU days | 21 (6-35) | 12 (4-18) | 0.2508 |
| 28-day mortality | 0 | 24 | 0.1336 |
| SOFA score | 7 (6-9) | 9 (8-12) | 0.0024 |
| APACHE II score | 12 (8-13) | 20 (14-25) | **0.0001** |
| Comorbidities: |  |  |  |
| Co-infections n (%) | 3 (18) | 9 (33) | 0.4295 |
| Cancer, n (%) | 0 (0) | 5 (18) | 0.0773 |
| Hypertension, n (%) | 8 (47) | 14 (50) | 0.8482 |
| Diabetes, n (%) | 2 (12) | 5 (18) | 0.6931 |
| Asthma, n (%) | 1 (6) | 3 (10) | 1.0000 |
| COPD, n (%) | 0 (0) | 1 (4) | 1.0000 |
| Obesity, n (%) | 3 (18) | 3 (10) | 0.6581 |
| Coronary artery disease, n (%) | 0 (0) | 4 (15) | 0.2812 |
| Chronic renal disease, n (%) | 0 (0) | 2 (7) | 0.7029 |
| Pregnancy, n (%) | 3 (18) | 0 (0) | **0.0160** |
| Secondary infections | 9 (53) | 12 (42) | 0.7522 |
| Treatment: |  |  |  |
| CRRT, n (%) | 0 (0) | 6 (21) | 0.1100 |
| ECMO, n (%) | 3 (18) | 8 (29) | 0.6390 |

**Abbreviations:** CCRT, continuous renal-replacement therapy; COPD, chronic obstructive pulmonary disease; ECMO, extra corporeal membrane oxygenation; ICU, intensive care unit;.

**Table 2.** Laboratory findings in the critically ill COVID-19 patients at the ICU admission

| Characteristic | survivors (n=17) | non-survivors (n=28) | p |
| --- | --- | --- | --- |
| White blood count [$\times 10^6/L$] | 12.9 (10.8-15.5) | 15.7 (11.6-18.9) | 0.3736 |
| Platelets [$\times 10^9/L$] | 287 (216-311) | 241 (178-332) | 0.2323 |
| APTT [seconds] | 33.1 (30.6-38.8) | 36.0 (30.6-42.9) | 0.3800 |
| INR | 1.10 (1.05-1.17) | 1.24 (1.15-1.37) | **0.0095** |
| D-dimers [$\mu g/ml$] | 1.80 (1.20-7.00) | 6.45 (1.90-23.70) | 0.0918 |
| C-reactive protein [$mg/L$] | 109.0 (67.0-149.0) | 110.0 (67.5-195.0) | 0.8149 |
| Procalcitonin [$ng/mL$] | 0.30 (0.11-0.43) | 0.51 (0.14-1.95) | 0.1528 |
| Interleukin-6 [$pg/ml$] | 75.85 (10.40-217.00) | 88.80 (40.05-1001.50) | 0.1939 |
| Glucose [$mg/dL$] | 136.0 (107.0-159.0) | 172.5 (127.0-214.0) | 0.1340 |
| Ferritin [$ng/ml$] | 698.5 (292.5-1563.5) | 1156.0 (568.0-2002.0) | 0.2089 |
| Lactate dehydrogenase [U/L] | 618.0 (480.0-709.0) | 583.0 (427.0-930.0) | 0.9906 |
| Galectin-1 [$pg/ml$] | 16424 (12957-22040) | 21760 (10759-29643) | 0.4329 |
| IL-1α [$pg/ml$] | 4.80 (4.80-11.97) | 5.46 (4.80-8.68) | 0.9905 |
| IL-1β [$pg/ml$] | 7.39 (2.69-11.46) | 8.08 (2.69-13.23) | 0.9252 |
| IL-1RA [$pg/ml$] | 43.11 (17.87-68.17) | 23.26 (10.71-87.75) | 0.7519 |
| IL-18 [$pg/ml$] | 57.78 (41.11-94.15) | 66.58 (24.14-96.61) | 0.9160 |
| ASC [$pg/ml$] | 0.00 (0.00-0.00) | 76.70 (18.70-134.700) | 1.0000 |
| GSDMD [$pg/ml$] | 0.02 (0.02-0.27) | 0.010 (0.06-0.12) | 0.5509 |

**Abbreviations:** APTT, activated partial thromboplastin time; ASC, apoptosis-associated speck- like containing a CARD; GSDMD, gasdermin D.

